# Sequencing SARS-CoV-2 in Slovakia: An Unofficial Genomic Surveillance Report

**DOI:** 10.1101/2021.07.13.21260431

**Authors:** Broňa Brejová, Viktória Hodorová, Kristína Boršová, Viktória Čabanová, Tomáš Szemes, Matej Mišík, Boris Klempa, Jozef Nosek, Tomáš Vinař

## Abstract

We present an unofficial SARS-CoV-2 genomic surveillance report from Slovakia based on approximately 3500 samples sequenced between March 2020 and May 2021. Early samples show multiple independent imports of SARS-CoV-2 from other countries. In Fall 2020, three virus variants (B.1.160, B.1.1.170, B.1.258) dominated as the number of cases increased. In November 2020, B.1.1.7 (alpha) variant was introduced in Slovakia and quickly became the most prevalent variant in the country (> 75% of new cases by early February 2021 and > 95% in mid-March).

## 1 Introduction

Genome sequence of the SARS-CoV-2 virus continually changes over time. The mutations eventually result in the emergence of new variants including those with higher infectivity, the ability to evade the immune system response, and causing milder or more severe clinical manifestations. It is therefore of utmost importance to constantly monitor the virus alterations by genome sequencing. Such monitoring provides a means for understanding the virus evolution and transmission, identification and characterisation of variants of concern (VoC), improvement of the tools for molecular diagnostics (e.g. RT-qPCR assays), as well as rapid adjustment of the health policy measures.

By the end of June 2021, global sequencing efforts yielded more than 2 millions genome sequences of the SARS-CoV-2 isolates from different geographical regions of the world. These sequences are available in public databases such as the GISAID initiative (http://www.gisaid.org/) and the European Nucleotide Archive (ENA, https://www.ebi.ac.uk/ena/) which allow rapid data sharing and provide robust resources for genomic epidemiology.

In this report, we summarize the results of the SARS-CoV-2 genomic surveillance in Slovakia during the first and second wave of the COVID-19 pandemic between March 2020 and May 2021.

## 2 Overview of Genomic Surveillance in Slovakia

The first SARS-CoV-2 samples in Slovakia were sequenced in March of 2020 by Comenius University Science Park using viral RNA amplified in VERO E6 cells and Illumina sequencing platform. Majority of samples in 2020 were sequenced using Oxford Nanopore MinION using the ARTIC PCR-tiling protocol originally developed for sequencing of Ebola and Zika virus samples (Quick et al., 2016, 2017), evaluating a variety of primer pools in the process (Brejova et al., 2021). In March 2021, a consortium of laboratories formed a genomic surveillance team that started routine sequencing of SARS-CoV-2 from clinical samples. The samples for sequencing are selected by the Public Health Authority of Slovakia and distributed to individual sequencing laboratories. Two laboratories (Public Health Authority and Comenius University Science Park) use Illumina sequencing, and Biomedical Centre of Slovak Academy of Sciences in collaboration with Comenius University in Bratislava is using MinION sequencing. All groups use variations of the ARTIC PCR-tiling protocol. Additional samples were sequenced at the Veterinary University in Zvolen and outside Slovakia (in Austria and Germany). Table 1 summarizes these efforts. Additional data for genomic surveillance have been obtained through differential qPCR testing designed to distinguish between common SARS-CoV-2 variants (Kováčová et al., 2021).

**Table 1:** Overview of sequence SARS-CoV-2 samples from Slovakia. The table shows numbers of samples submitted to GISAID with collection dates between March 2020 and March 2021. The numbers in parenthesis indicate median response time in days (from collection to submission).

|  |  |  |  |  |  |
| --- | --- | --- | --- | --- | --- |
| BMC + Comenius University - | 1 (196) | 8 (175) | 11 (37) | 63 (39) | 591 (14) |
| CeMM Austria - | - | - | - | 60 (76) | 88 (63) |
| Charite Berlin - | 2 (162) | - | - | - | - |
| Comenius University Science Park - | 4 (20) | - | - | 42 (108) | 1086 (27) |
| Public Health Authority - | - | - | - | - | 193 (79) |
| Veterinary Institute Zvolen - | - | 4 (386) | 4 (291) | 5 (181) | 7 (78) |
|  | Q1 2020 | Q2 2020 | Q3 2020 | Q4 2020 | Q1 2021 |

## 3 Early Cases (March-June 2020)

The first case of SARS-CoV-2 infection in Slovakia has been documented in Kostolište (Malacky district in western Slovakia) on March 6, 2020, in a 52 year old man (The Slovak Spectator, 2020). The next day, two members of his family were tested positive for the infection (SK-BMC1), including his son who returned from Venice, Italy, and presumably got infected while traveling (Úrad verejného zdravotníctva SR, 2020). In the following days, additional cases were confirmed in unrelated persons in Bratislava, Košice, and Martin. Rapid introduction of prevention and control measures including the diagnostics based on real-time quantitative polymerase chain reaction (RT-qPCR), contact tracing, national lockdown, and quarantine for travelers led to substantial reduction of the virus spreading in the country.

The phylogeny of 19 early samples collected between March and June 2020 (Table 2, Figure 1) suggests at least six unrelated import events, likely through routine international travel. The closest matches from GISAID database based on the Jaccard index include samples from France (SK-BMC5), Netherlands (SK-BMC6), Ireland (UKBA-204), Scandinavia (60007-VHU_185), Serbia (UKBA-209), Ukraine (UKBA-210), and the United Kingdom (UKBA-207, UKBA-208). Note that these are locations where particular mutation combinations were common; sampled individuals could have contracted COVID-19 elsewhere.

**Table 2:** Early cases documented in Slovakia with collection dates between March and June 2020.

| ID | Collection date | Pangolin lineage | Location | Mut. count |
| --- | --- | --- | --- | --- |
| hCoV-19/Slovakia/ChVir-1996/2020 | 2020-03 | B.1 | Bratislava | 6 |
| hCoV-19/Slovakia/ChVir-1998/2020 | 2020-03 | B.1 | Bratislava | 7 |
| hCoV-19/Slovakia/SK-BMC1/2020 | 2020-03-06 | B.1 | Kostolište | 6 |
| hCoV-19/Slovakia/SK-BMC5/2020 | 2020-03-06 | B.1 | Košice | 7 |
| hCoV-19/Slovakia/SK-BMC2/2020 | 2020-03-07 | B.1 | Bratislava | 7 |
| hCoV-19/Slovakia/SK-BMC6/2020 | 2020-03-08 | B.11 | Martin | 2 |
| hCoV-19/Slovakia/UKBA-212/2020 | 2020-03-31 | B.1.1 | Bratislava | 7 |
| hCoV-19/Slovakia/UKBA-101/2020 | 2020-04-03 | B.1 | Bratislava | 11 |
| hCoV-19/Slovakia/UKBA-203/2020 | 2020-04-06 | B.1.1 | Žehra | 10 |
| hCoV-19/Slovakia/UKBA-204/2020 | 2020-04-06 | B.1.1 | Bratislava | 11 |
| hCoV-19/Slovakia/UKBA-205/2020 | 2020-04-13 | B.1.1 | Žehra | 11 |
| hCoV-19/Slovakia/60007-VHU_185/2020 | 2020-04-15 | B.1 | Hencovce | 9 |
| hCoV-19/Slovakia/60020-VHU_1250/2020 | 2020-04-24 | B.1.1 | Martin | 8 |
| hCoV-19/Slovakia/60023-VHU1453_2020/2020 | 2020-04-27 | B.40 | Petrovany | 12 |
| hCoV-19/Slovakia/UKBA-207/2020 | 2020-04-29 | B.1.1 | Pezinok | 11 |
| hCoV-19/Slovakia/UKBA-208/2020 | 2020-05-06 | B.1.1 | Pezinok | 8 |
| hCoV-19/Slovakia/60056-VHU_4065/2020 | 2020-05-15 | B.1.1 | Martin | 8 |
| hCoV-19/Slovakia/UKBA-209/2020 | 2020-06-30 | B.1.1.70 | Bratislava | 16 |
| hCoV-19/Slovakia/UKBA-210/2020 | 2020-06-30 | B.1.131 | Bratislava | 12 |

**Figure 1:**
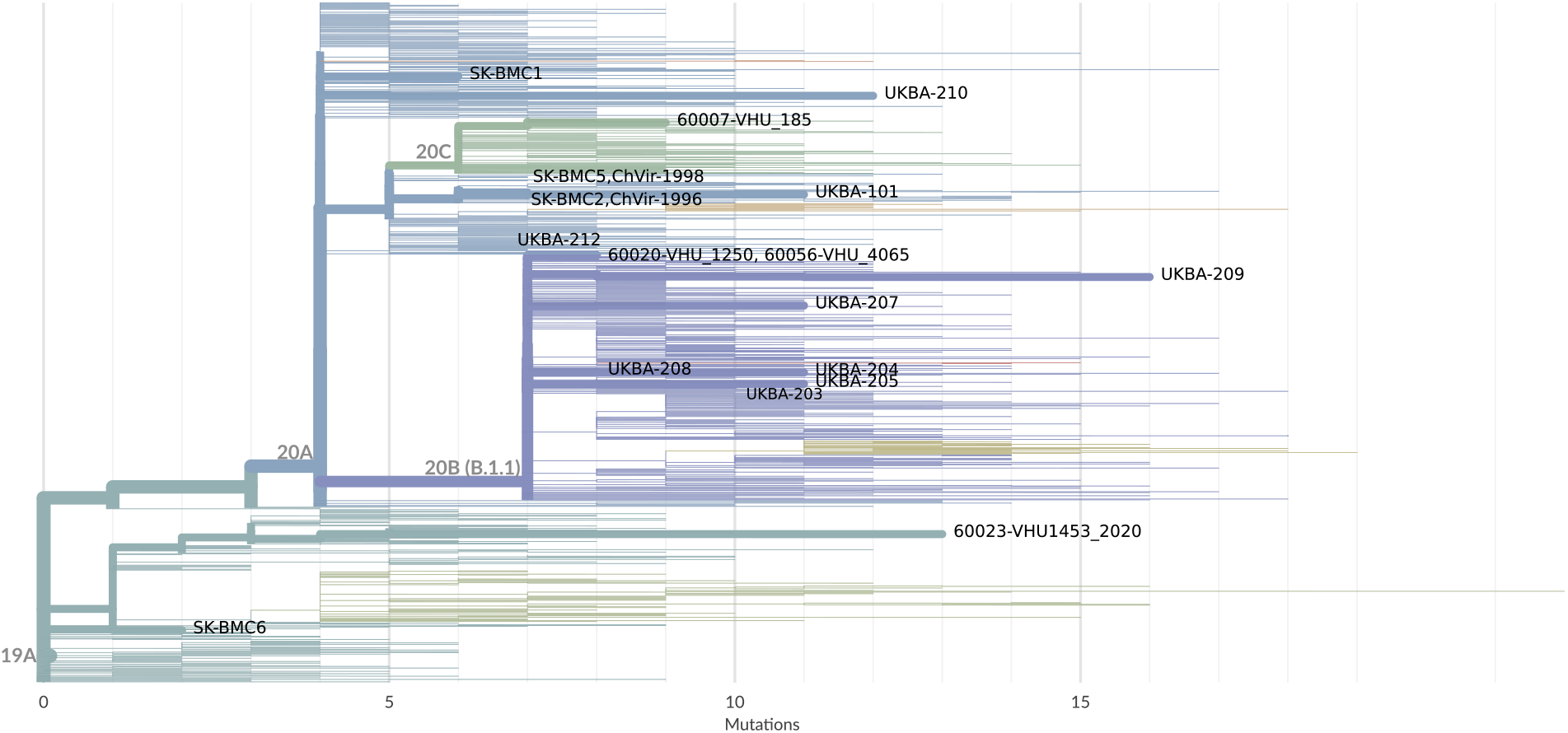
Phylogeny of early cases from Slovakia on background of randomly chosen samples from GISAID.

Three samples (SK-BMC2, UKBA-205, UKBA-101) were related to the fallout from a documented superspreading event at a medical conference in Boston, MA at the end of February (Lemieux et al., 2021). Sample UKBA-212 is identical to 3757 additional samples from all over the world (including United Kingdom, United States, Portugal, and Italy), which combined an earlier mutation Spike:D614G, which increases the infectivity and stability of virions, leading to higher viral loads and competitive fitness (Plante et al., 2021), with mutations in nucleoprotein N:R203K and N:G204R. Large number of identical samples indicates very fast spread perhaps following a superspreading event, and the group has become a foundation for evolution of B.1.1 lineage; see also analysis of Austrian samples (Popa et al., 2020) and global analysis of early SARS-CoV-2 lineages (Gómez-Carballa et al., 2020). In Slovakia, nine early samples are classified to the B.1.1 lineage (including sublineage B.1.1.70).

## 4 Rise in the Fall of 2020

The release of restrictions during the summer 2020 increased the number of imported cases, which has been followed by community transmission. The quick spread of the infection throughout the country with about 5.5 million inhabitants raised the cumulative number of infected people to 267147 as of December 31, 2020. While in many European countries, B.1.177 (EU1) lineage has become dominant in Fall of 2020 (Hodcroft et al., 2021), different three lineages appear to have achieved a substantial prevalence in Slovakia between September and November 2020, namely B.1.1.170, B.1.160 (EU2), and B.1.258 (Figure 2).

**Figure 2:**
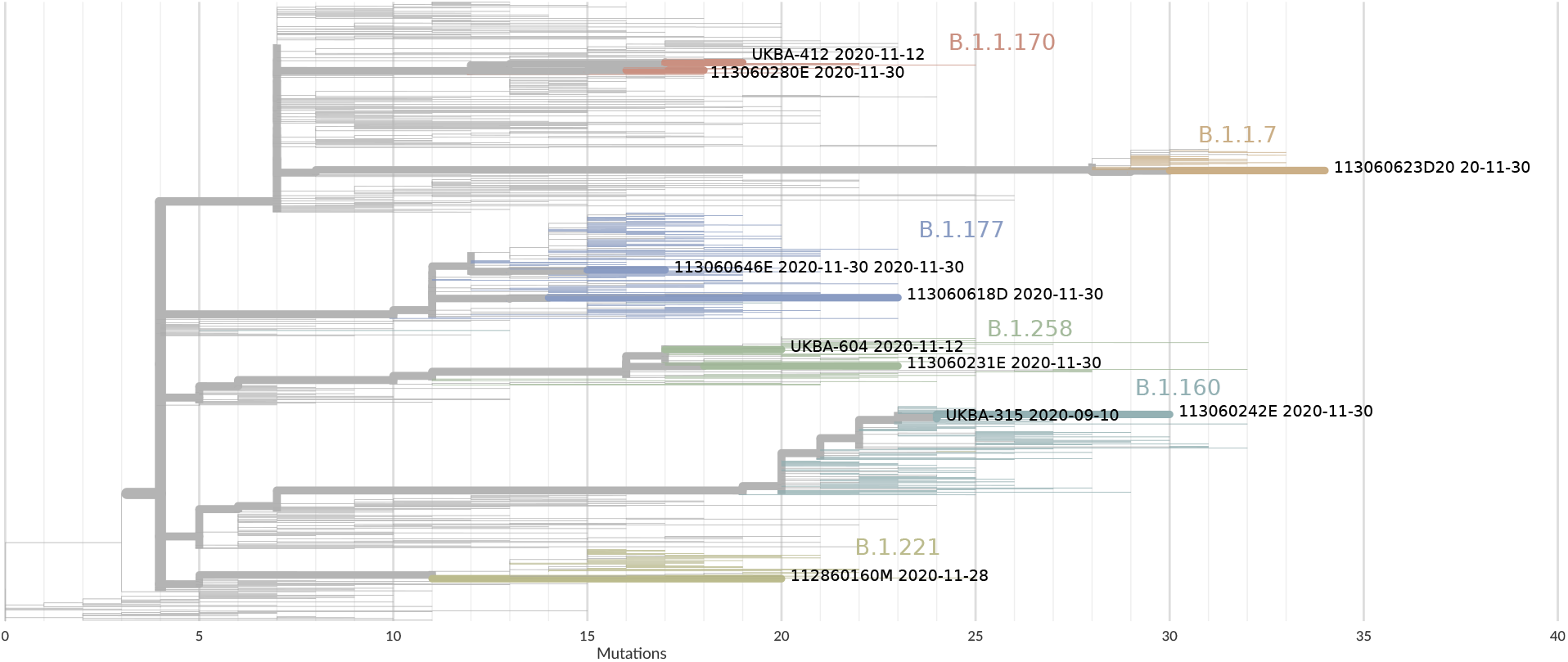
Selected samples from Slovakia collected between September and December 2020 on the background of randomly chosen samples from GISAID. Lineages of interest, namely B.1.1.170, B.1.160, B.1.1.7, B.1.221, and B.1.258, are overrepresented among the background samples.

B.1.1.170 lineage is characterized by mutation P822H (C5184A) in NSP3 peptidase C16 domain required for proteolytic processing of replicase polyprotein. This lineage has been observed in other countries in high numbers, including Denmark, Germany, and United Kingdom; however, in neither of these countries B.1.1.170 represented a significant percentage of cases in the population (Figure 3). Instead, the high absolute number of B.1.1.170 cases reflects the large scale of the sequencing programs in these countries.

**Figure 3:**
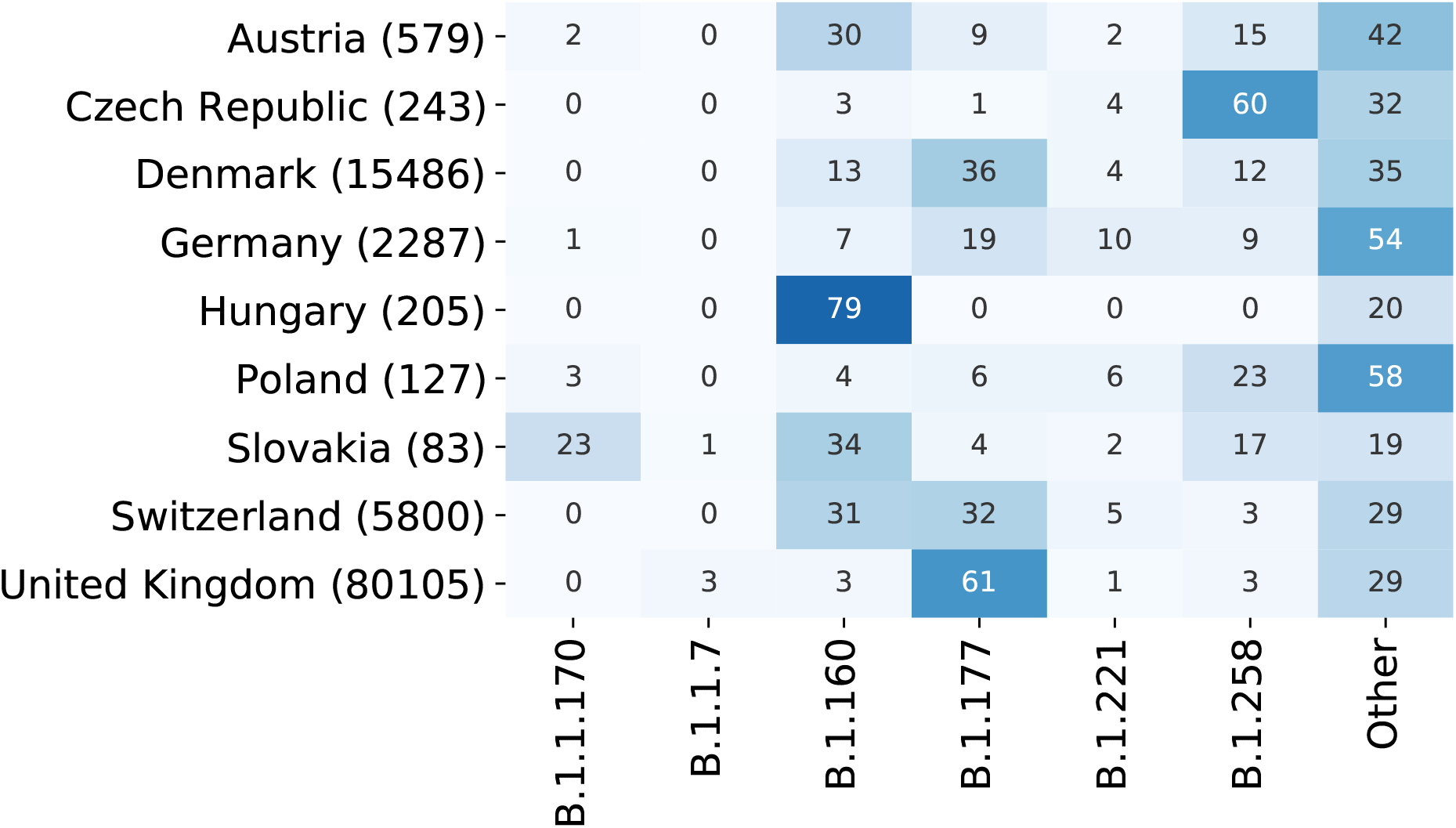
Prevalence of SARS-CoV-2 lineages in sequenced samples collected between September and November 2020. The number in parenthesis indicates the number of sequenced samples deposited in GISAID with collection dates within this time period. The count for each studied lineage includes also all its sublineages.

B.1.160 (EU2) lineage is characterized by amino acid substitutions M234I and A376T in N, M324I (G9526T) in NSP4, A176S, V767L, K1141R, E1184D in ORF1b, and S477N in S (Fournier et al., 2021). The position 477 in the receptor binding motif of the Spike protein plays a crucial role in the interaction with the human receptor ACE2 (Singh et al., 2021) and the mutant has been shown to increase the infectivity (Chen et al., 2020b); the same mutation has also emerged in an unrelated outbreak in Australia (Chen et al., 2020a). Besides Slovakia, the B.1.160 was highly prevalent in Hungary, Austria, and Switzerland (Figure 3).

B.1.258 lineage harbours Spike protein receptor binding domain mutation N439K, shown to enhance the binding affinity of the Spike protein to human immune response (Thomson et al., 2021), and the sublineages prevalent in Central Europe combine this mutation with ΔH69/ΔV70 deletion, facilitating escape from immune response (Kemp et al., 2021), which is also one of the characteristic mutations of B.1.1.7 (alpha) lineage emerging later. Lineage B.1.258 likely originated in Switzerland (Brejová et al., 2021), and spread mainly in Central European countries, including Czech Republic, Slovakia, Poland, Austria, and Germany (Figure 3).

Interestingly, B.1.177 (EU1) lineage, wide-spread across many European countries during Fall of 2020 does not show any evidence of increased transmissibility. It seems to have become dominant simply by repeated introduction to respective countries by summertime travellers, undermining local efforts to keep SARS-CoV-2 cases low (Hodcroft et al., 2021). In Slovakia, this lineage was discovered only at the end of November 2020 and sporadically appeared in sequencing samples since then.

The mutation rate of B.1.177 and B.1.1.170 seems to be close to the background mutation rate of 1.4 mutations per month, obtained by linear regression from GISAID samples excluding samples belonging to the known variants characterized by a significant increase in numbers of mutations (Figure 4). In contrast, lineages B.1.160 (EU2) and B.1.258 show an interesting evolutionary pattern, where a branch leading to the lineage in the phylogenetic tree is associated with a surge in mutations. After this initial surge, the evolutionary rate stabilizes again at the mutation rate close to the background mutation rate (Figure 4). Such unusual genetic divergence has also been observed in B.1.1.7 (Rambaut et al., 2020) and other VoCs (Figure 4), and is likely indicative of positive selection. One possible mechanism is a selective pressure upon the within-patient virus population in immunodeficient or immunosuppressed chronically infected patients treated with convalescent plasma and antiviral drugs (Choi et al., 2020; Avanzato et al., 2020; Kemp et al., 2021).

**Figure 4:**
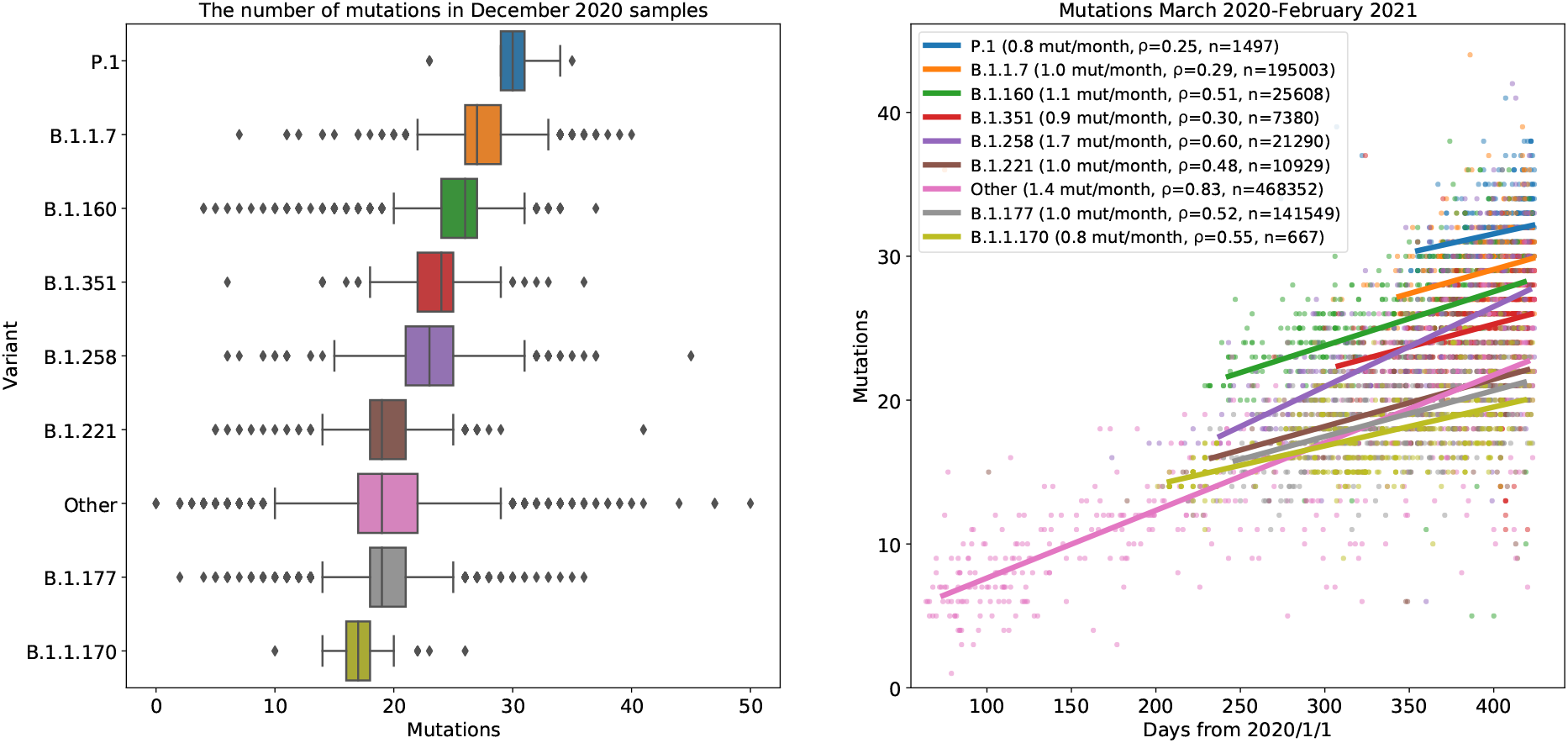
Evolution of SARS-CoV-2 lineages over time. (left) The number of mutations compared to the reference virus sequence in samples collected in December 2020. The samples were split according to identified lineage. (right) Accumulation of mutations in individual variants over time. Linear regression shows the base mutation rate of 1.4 substitutions per month (line “other”, Pearson correlation 0.83), and mutation rates in selected lineages varying between 0.8 mutations per month in P.1 and 1.7 mutations per month in B.1.258.

While in Spring of 2020, Slovakia was one of the countries with the best response to the pandemic situation (Serhan, 2020), the Fall was characterized by a steep rise in cases. Besides slow and inconsistent response of government institutions to the worsening situation, the genomic surveillance from that period highlights Slovakia’s position at the crossroads of Central Europe, which likely led to importation of new variants from neighbouring countries. Combined effect of these variants, some of which share evolutionary characteristics with later identified variants of concerns, likely contributed to worsening of the pandemic situation.

## 5 Introduction of B.1.1.7

Variant B.1.1.7 (alpha) was first observed on September 20, 2020 in Kent, United Kingdom (Rambaut et al., 2020) and quickly spread throughout the United Kingdom and the world. Out of unusually large number of mutations in the spike protein, substitution N501Y in the receptor-binding domain has been identified to increase the binding affinity to human ACE2 receptors, the ΔH69/ΔV70 deletion increases infectivity and mediates cell-cell fusions, ΔY144 is localized in an antibody supersite epitope, and P681H is adjacent to biologically significant furin cleavage site (Meng et al., 2021; Gupta, 2021).

The first documented case in Slovakia was collected in Bratislava on November 30, 2020. Due to the low volume of sequencing at the time, it is difficult to estimate the prevalence and the spread of the B.1.1.7 variant over time. However, antigen mass testing in the city of Trenčín in Western Slovakia on December 19-20, followed by qPCR re-testing on a voluntary basis, yielded 148 PCR-positive samples collected on December 22 (Mesto Trenčín, 2021), out of which 122 were later re-tested using tests designed to differentiate between B.1.1.7 and B.1.258 (both carrying ΔH69/ΔV70), and variants that do not harbour this deletion (Kováčová et al., 2021). Out of these, 4 samples (or 3.3%) were identified as B.1.1.7, and this was also confirmed by sequencing of selected samples (Brejová et al., 2021). The fraction of B.1.1.7 cases in the city of Trenčín has quickly risen to 76.1% in as little as 42 days, according to the nation-wide differential qPCR testing on February 3, 2021 (see https://github.com/Institut-Zdravotnych-Analyz/covid19-data).

Interestingly, the increase from 3.3% to 76.1% in 42 days is fast in comparison with other countries. Among the countries with high number of sequenced samples, Denmark took 48 days to rise from ≈ 3.3% to ≈ 76.1% (between December 29 and February 15), in United Kingdom and Switzerland, a similar rise in the fraction of cases took 61 days (October 31-December 31) and 62 days (December 19-February 19) respectively, and in Germany it took even longer (77 days between December 17 and March 4). While it is difficult to speculate on why the B.1.1.7 was able to achieve domination in Slovakia so quickly, it is worthwhile to point out that during this period, Slovakia was under various forms of nation-wide lockdown. It has been demonstrated that effectiveness of lockdown measures differs between old variants and B.1.1.7 (Vöhringer et al., 2020). Also, a fatigue from following the rules and inconsistencies in the government imposed interventions likely caused people to selectively choose to follow certain rules while rejecting others, based mostly on their own experience. Such an approach may have increased the lineage-based differences in effectiveness of mitigation.

One possible explanation for fast spread of B.1.1.7 in Slovakia are repeated imports by workers and students visiting home during the Christmas holidays. In fact, one of the first B.1.1.7 outbreaks detected in Slovakia was in a marginalized community in the Eastern Slovakia (Pavlovce nad Uhom), where the link to travel from the United Kingdom was clearly established. However, we show below that this may not be the main factor.

Interestingly, 74% of B.1.1.7 sequenced cases collected in Slovakia between November 2020 and May 2021, form a separate clade in the phylogenetic tree (called B.1.1.7ce for the purpose of this paper), characterized by mutations C5944T and G28884C (Figure 5). While the first of these is silent, the second causes R204P substitution in the nucleoprotein IDR2 region. Note that this site was mutated from G to R at the base of lineage B.1.1. The G28884C mutation also extends an existing span of three consecutive mutations at positions 28881-28883 compared to the reference, this region being characterized as a mutational hotspot of the N protein (Azad, 2021). Additional mutations A28095T (ORF8:K68*; mutations in ORF8 being potentially linked to immune evasion (Zhang et al., 2021) and T15096C likely happened after the emergence of B.1.1.7, but prior to the characteristic mutations C5944T and G28884C. The earliest case in GISAID belonging to B.1.1.7ce sublineage has been collected in Switzerland on November 9, 2020, the early cases from Bratislava (November 30, 2020) and Trenčín (December 22, 2020) also belong to the ce sublineage. Besides Slovakia, sublineage B.1.1.7ce represents 91% of sequenced B.1.1.7 samples from the Czech Republic, 69% in Hungary, and 57% in Austria. (Surprisingly Colombia and French Guiana also show over 40% cases of B.1.1.7 belonging to the B.1.1.7ce sublineage, but the number of sequenced genomes are quite small, and thus the sample may not be representative). In contrast, B.1.1.7ce constitutes only 0.3% of B.1.1.7 samples in the United Kingdom.

**Figure 5:**
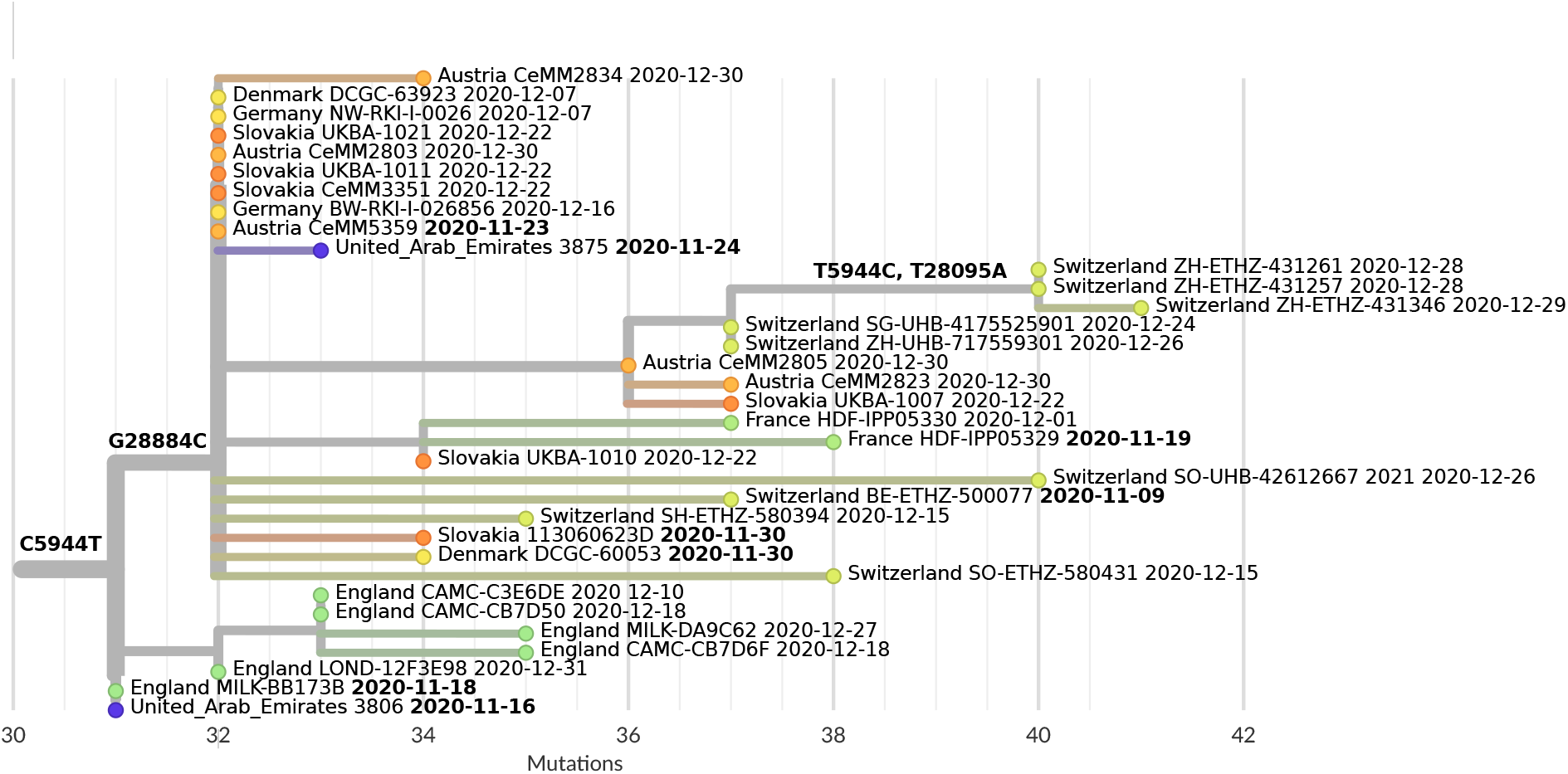
Early cases of B.1.1.7ce clade. Note that this particular reconstruction includes apparent back mutations at positions 5944 and 28095. However, these may be caused by data processing artefacts as these positions are within the ARTIC v3 primer binding sites, which may render them invisible to some computing pipelines typically used for processing SARS-CoV-2 sequencing data.

Phylogenetic tree reconstruction from a randomly selected subset of B.1.1.7 cases from Slovakia, Czech Republic, and Austria shows a large radiation at the base of B.1.1.7ce clade, which suggests a rapid spread of this clade before further mutations had a chance to accumulate. High percentage of these samples in Central European countries contradicts the theory of repeated introduction by independent travelers from the United Kingdom (with only 0.3% of B.1.1.7ce cases out of all B.1.1.7 cases), and instead suggests a fast community spread directly within Central Europe. This theory is further supported by the data from four screenings by differential qPCR tests performed between February 3 and March 17, 2021, initially showing high percentage of B.1.1.7 cases in Western Slovakia and spreading over time to the eastern parts of the country (Figure 7; Kováčová et al. (2021)).

Lineage B.1.1.7 constitutes 96.7% of 3184 GISAID samples collected in Slovakia between February and May 2021. Other lineages previously present in Slovakia, such as B.1.258, B.1.160, B.1.1.170, B.1.177, constitute 54 samples in total (1.7%). Other lineages occur sporadically and in many cases have been linked directly to international travel with only a limited community spread. Lineage B.1.351 (VoC Beta) was found in 27 samples. Finally, 25 samples belong to 12 additional lineages, including ECDC variants of interest (European Centre for Disease Prevention and Control, 2021a) B.1.617.1 (Kappa) and B.1.621.

## 6 Discussion and Conclusions

Throughout 2020, genomic surveillance of COVID-19 pandemic in Slovakia consisted almost exclusively from uncoordinated activities of individual researchers, which has resulted in highly uneven sequencing coverage, both in time and regionally. Nevertheless, the information collected has provided a unique insight into a progression of COVID-19 pandemic in Slovakia and allowed us to identify both common and unique trends compared to the neighbouring countries. The situation changed in March 2021 with the establishment of coordinated efforts involving the Public Health Authority, Comenius University, and Biomedical Center of Slovak Academy of Sciences. Since then, the Public Health Authority has been selecting and distributing positively tested PCR samples to individual labs for sequencing and the number of sequenced samples typically exceeded 500 samples per week in June 2021.

Yet, there is space for improvements. A major problem currently lies with the logistics, where samples are delivered to the sequencing labs two weeks or longer after their collection. Depending on the laboratory and sequencing technology used, the time from receiving samples to sequencing results can be as short as two days or as long as one week. The information obtained through sequencing is thus much delayed and has only a limited value for treatment, epidemiological response, or as the basis for rapid public policy decisions. Examples from Denmark, United Kingdom, Netherlands, and other countries (see e.g. (Oude Munnink et al., 2020)) show that these logistic issues can be solved and the response time can be decreased dramatically. In fact, governments in these countries routinely use the information obtained through sequencing to fine-tune the pandemic mitigation measures.

According to the recommendations from the ECDC (European Centre for Disease Prevention and Control, 2021b), in choosing samples for sequencing, the priority should be given to the representative sampling for the purpose of surveillance of emerging variants (even those that are not yet characterized as VoCs). This can be combined with targeted monitoring of outbreaks, vaccine escape and reinfection, long-term persistent infections, monitoring of travel, etc. For the purpose of data analysis, it is essential that the reasons for choosing a particular sample for sequencing is known to the researchers analyzing the data; yet this information is not provided by the Public Health Authority and data analysts have no influence in developing the sampling strategy. Based on recently improving epidemiological situation, there is currently an ambition to sequence all samples with sufficient viral load. Nevertheless, this issue is likely to reappear once the situation worsens and the selection of samples is again necessary.

While some of the analyses requiring integration of epidemiology and genomics data are conceptually straightforward (such as monitoring the prevalence of lineages over time), other tasks, such as recognizing mutations spreading due to a selective advantage rather than a random drift, are much more involved and require complex bioinformatics and modeling expertise (see, e.g. (Vöhringer et al., 2020)). At present, we are not aware of any plans of establishing a team that would have such an expertise, enough redundancy to perform such analyses regularly, unobstructed access to all necessary data, and regular communication with experts elsewhere on these matters.

## 7 Methods

SARS-CoV-2 genomic sequences and their metadata (including date of collection and submission, country, and Pangolin lineage) were downloaded from GISAID (Shu and McCauley, 2017) on June 18, 2021. The database contained 2,012,564 sequences. Out of these, we have used 1,950,347 sequences with fewer than 5kbp of missing sequence.

The presence of individual substitutions was ascertained by mapping individual genomes to the reference hCoV-19/Wuhan/Hu-1/2019 by minimap2 (Li, 2018) and then formatting the result into a multiple alignment in reference sequence coordinates by gofasta tool (Jackson, 2020). To count mutations for Figure 4, each continuous stretch of mutated bases was counted as a single mutation to mitigate impact of occasional local misalignments. In this figure, only sequences with fully specified date, with at most 1kb of missing sequence and at most 50 mutations were used. Sequences with more than 50 mutations were rare in the displayed period. All sublineages of each displayed lineage were included within this lineage. All samples were used to estimate linear regression, but at most 500 samples (randomly selected) are displayed in the plot per lineage.

Phylogenetic trees were created by Augur and visualized by Auspice (Hadfield et al. 2018; Sagulenko, Puller, and Neher 2018). Tree in Figure 1 contains all samples collected in Slovakia before July 2020. As a background, we have selected 10% of samples from other countries sequenced before March 2020 and 1% of samples selected between March and June 2020 (inclusive). We have removed all samples with more than 25 mutations compared to the reference as likely metadata errors (samples from early 2021 mistakenly marked as 2020). Finally the set of 1830 background samples was reduced to 1199 by removing samples that differed by the presence or absence of at most one mutation from some older sample. The tree in Figure 2 highlights manually selected representative Slovak samples from lineages described in the text. The background sequences were randomly selected with probably 0.2% from samples sequenced before the end of November 2020 with at most 40 mutations compared to the reference. To give a better context for the selected samples, some lineages were overrepresented in the background set. Namely, 10% of samples from B.1.1.170 were added, as well as 1% of samples from B.1.160, B.1.1.7, B.1.221, B.1.258; in both cases using only samples from September to November 2020. Again, the background set was reduced by the removal of samples differing by at most one mutation. The tree in Figure 5 is a clade selected from a bigger tree, which contained three outgroup sequences (reference hCoV-19/Wuhan/Hu-1/2019, an early B.1.1 sample hCoV-19/Slovakia/UKBA-212/2020, and an early B.1.1.7 sample hCoV-19/England/MILK-9E05B3/2020), 968 B.1.1.7 samples collected in 2020 and containing the A28095T mutation (out of all 2990 samples satisfying these criteria, we have again filtered out nearly identical sequences). We have also added all B.1.1.7 sequences from 2020 that contain at least one of the mutations G28884C and C5944T characteristic for B.1.1.7ce clade. We have excluded 8 sequences that appeared as outliers markedly different from other sequences, possibly due to recombination or technical errors. Finally, the tree in Figure 6 contains a selection of B.1.1.7 sequences from Austria, Czech Republic and Slovakia collected between September 2020 and May 2021, excluding samples marked as environmental as well as sequences from Slovakia lacking G28882A mutation. Many samples without this mutation were creating a spurious clade; we believe that the lack of this mutation is due to technical problems with calling the four successive variants 28881-28884 in B.1.1.7ce using very short Illumina reads. From the remaining samples, we have taken 25% of sequences from Slovakia and Austria and 14% from the Czech Republic. After again filtering out nearly identical sequences in each country separately, we were left with 361 Austrian, 382 Czech and 371 Slovak sequences. We added the reference hCoV-19/Wuhan/Hu-1/2019 as an outgroup and removed three outliers.

**Figure 6:**
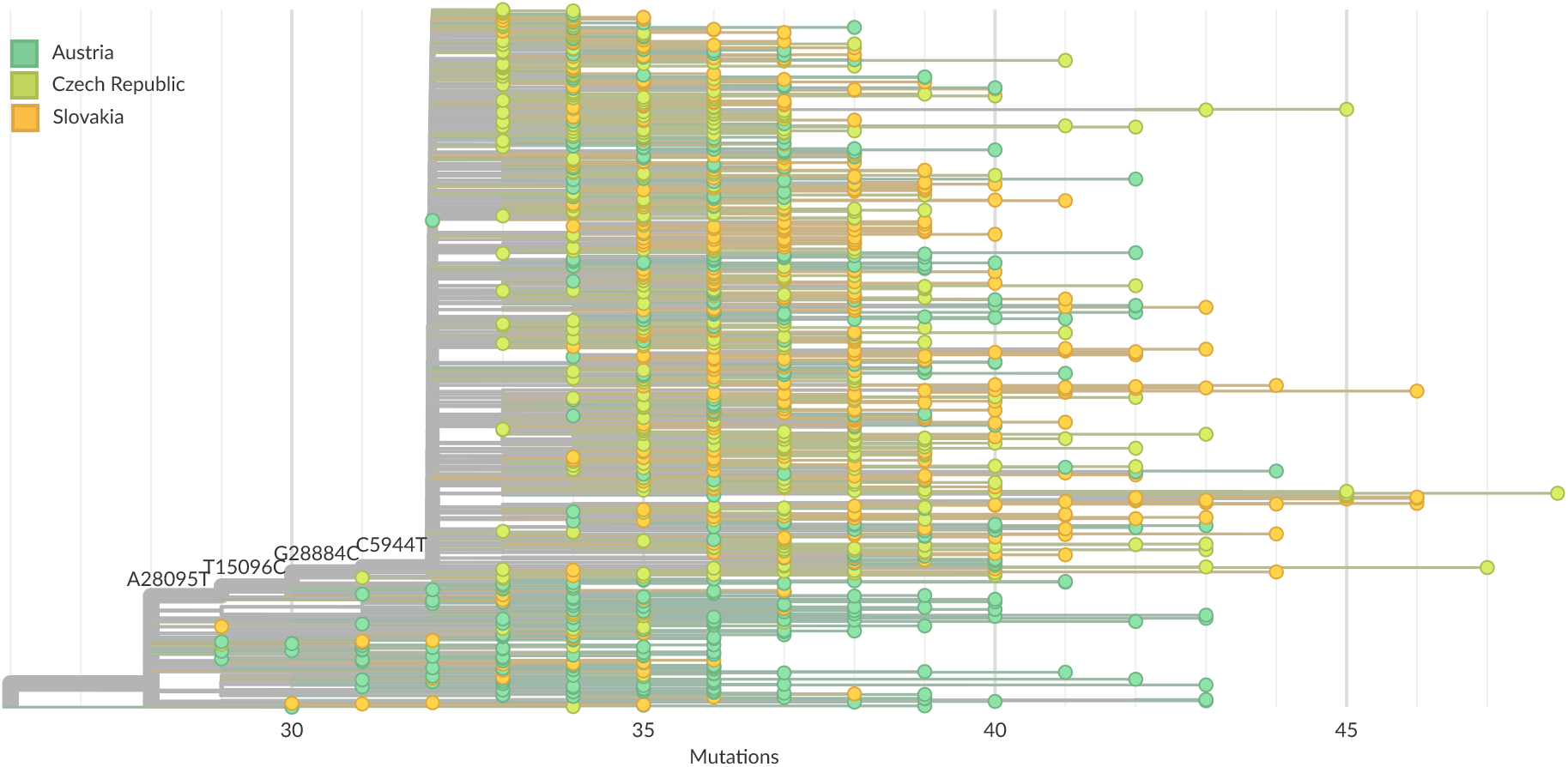
A randomly selected subset of B.1.1.7 samples from Slovakia, Czech Republic and Austria.

**Figure 7:**
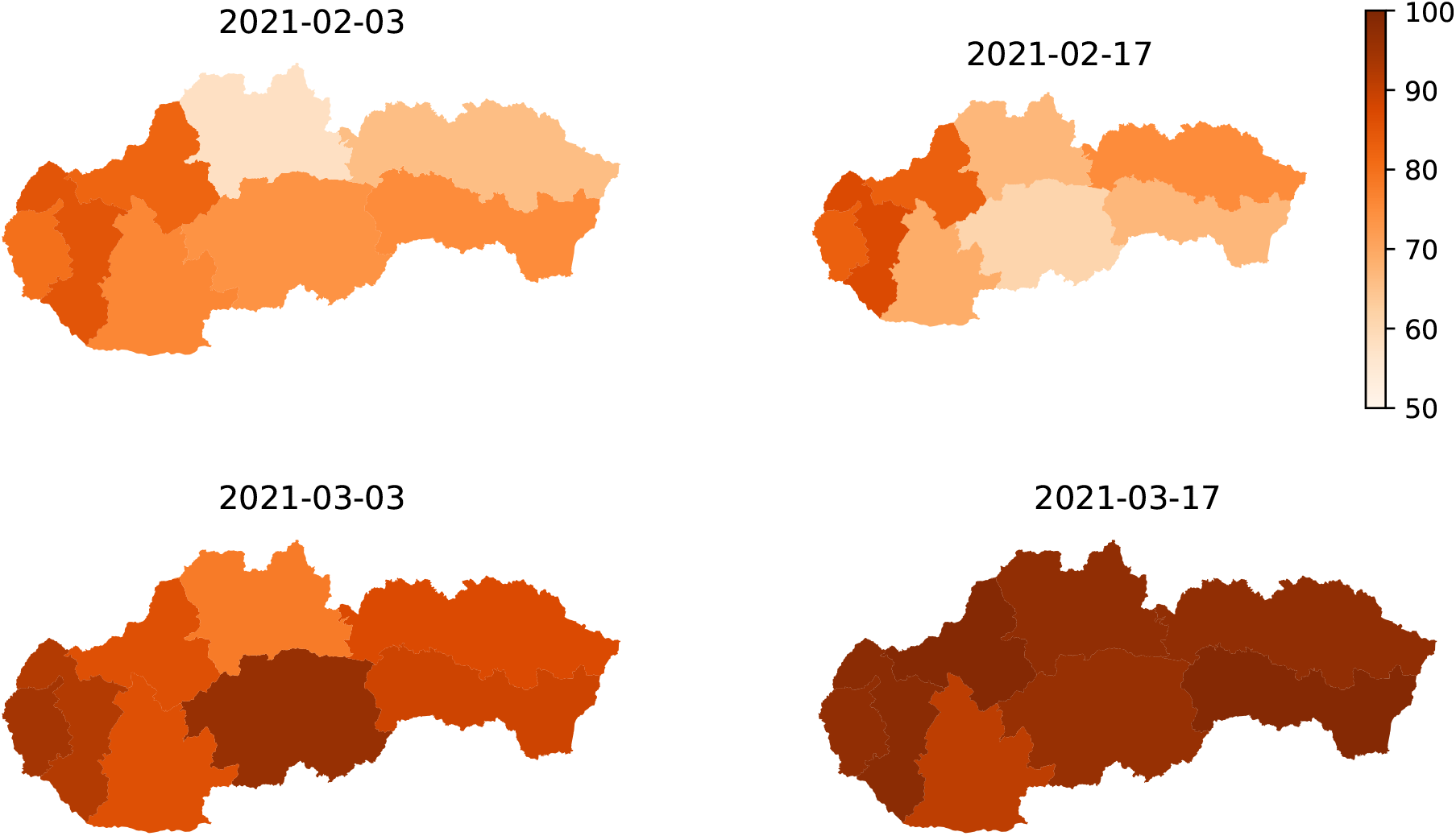
Percentage of B.1.1.7 among samples positively tested by qPCR in different regions of Slovakia. All samples positively tested on these dates were re-tested with differential qPCR tests designed to distinguish B.1.1.7, B.1.258, and variants that do not harbour ΔH69/ΔV70 deletion (data: Institute of Health Analyses).

## Supporting information

GISAID acknowledgements

## Data Availability

All data are available through GISAID database.

https://gisaid.org

## Ethical statement

The study has been approved by the Ethics committee of Biomedical Research Center of the Slovak Academy of Sciences, Bratislava, Slovakia (Ethics committee statement No. EK/BmV-02/2020).

## Acknowledgements

This research has been supported by the Operational Program Integrated Infrastructure project ITMS:313011ATL7 “Pangenomics for personalized clinical management of infected persons based on identified viral genome and human exome” (90%) co-financed by the European Regional Development Fund. The research was also supported by a grant from VEGA 1/0458/18 to TV (10%).

We gratefully acknowledge the authors from the originating laboratories responsible for obtaining the specimens, as well as the submitting laboratories where the genome data were generated and shared via GISAID (https://www.gisaid.org/), on which this research is based.

