## Supplementary material for "Sequencing SARS-CoV-2 in Slovakia: An Unofficial Genomic Surveillance Report": GISAID acknowledgements

| Originating laboratory | Submitting laboratory | Authors | GISAID EPI ISL |
|---|---|---|---|
| Public Health Authority of the Slovak Republic, Department of Medical Microbiology | Charite Universitatsmedizin Berlin, Institute of Virology | Victor M Corman, Terry Jones, Jörn Beheim-Schwarzbach, Barbara Muehlemann, Talitha Veith, Julia Schneider, Mgr. Edita Staronova, Christian Drosten | EPI_ISL_516988, EPI_ISL_516989 |
| Regional Authority of Public Health Banská Bystrica | Veterinary institute in Zvolen Slovakia | Dirbáková Z., Maďarová L., Mancoš M., Strhársky J., Sujová S., Mokryšová S., Tinák M., Mojžiš M. | EPI_ISL_2254725, EPI_ISL_2254726, EPI_ISL_2255181 |
| Public Health Authority of the Slovak Republic, Bratislava | Faculty of Natural Sciences, Comenius University in Bratislava | Dominika Fričová, Viktória Hodorová, Kristína Boršová, Broňa Brejová, Viktória Čabanová, Sabina Fumačová Havlíková, Juraj Kopáček, Martina Ličková, Ľubomíra Lukáčiková, Martina Neboháčová, Monika Sláviková, Edita Staroňová, Elena Tichá, Tomáš Vinař, Boris Klempa, Jozef Nosek | EPI_ISL_572329 |
| Hospital | National Reference Center for Viruses of Respiratory Infections, Institut Pasteur, Paris | Marion Barbet, Sylvie Behillil, Méline Bizard, Angela Brisebarre, Camille Capel, Louise Lefrançois, Etienne Simon-Lorière, Vincent Enouf, Maud Vanpeene, Sylvie van der Werf, Hecquet Denise | EPI_ISL_1381145, EPI_ISL_1381146 |
| Institute of Virology, Biomedical Research Center of the Slovak Academy of Sciences, Bratislava; Public Health Authority of the Slovak Republic, Bratislava | Institute of Virology, Biomedical Research Center of the Slovak Academy of Sciences, Bratislava; Comenius University Science Park, Bratislava | Monika Slávikova, Martina Ličková, Sabina Fumačová Havlíková, Juraj Koči, Juraj Kopáček, Elena Tichá, Edita Staroňová, Jaroslav Budiš, Werner Krampl, Miroslav Böhmer, Diana Rusňáková, Tomáš Szemeš, Boris Klempa | EPI_ISL_417877, EPI_ISL_417878, EPI_ISL_417879, EPI_ISL_417880 |

| | | | |
|---|---|---|---|
| Viollier AG | Department of Biosystems Science and Engineering, ETH Zürich | Chaoran Chen, Sarah Nadeau, Catharine Aquino, Ivan Topolsky, Philipp Jablonski, Lara Fuhrmann, David Dreifuss, Katharina Jahn, Andreia Cabral de Gouvea, Maria Domenica Moccia, Simon Grüter, Timothy Sykes, Lennart Opitz, Griffin White, Laura Neff, Doris Popovic, Andrea Patrignani, Jay Tracy, Ralph Schlapbach, Christiane Beckmann, Maurice Redondo, Olivier Kobel, Christoph Noppen, Sophie Seidel, Noemie Santamaria de Souza, Niko Beerenwinkel, Tanja Stadler | EPI_ISL_1750432, EPI_ISL_1750710 |
| Institute of Virology, Biomedical Research Center of the Slovak Academy of Sciences, Bratislava | Faculty of Natural Sciences, Comenius University, Bratislava | Viktória Čabanová, Kristína Boršová, Broňa Brejová, Viktória Hodorová, Sabina Fumačová Havlíková, Juraj Kopáček, Martina Ličková, Ľubomíra Lukáčiková, Martina Neboháčová, Monika Slávikovà, Tomáš Vinař, Jozef Nosek, Boris Klempa | EPI_ISL_903989, EPI_ISL_904000 |
| Labormedizinisches Zentrum Dr Risch | University Hospital Basel, Clinical Bacteriology | Tim Roloff, Madlen Stange, Helena MB Seth-Smith, Alfredo Mari, Karoline Leuzinger, Julia Bielicki, Nadia Wohlwend,Martin Risch, Lorenz Risch, Manuel Battegay, Hans Hirsch, Adrian Egli | EPI_ISL_861861, EPI_ISL_861863 |
| University College London, Great Ormond Street Hospital for Children NHS Foundation Trust, Imperial College Healthcare NHS Trust | COVID-19 Genomics UK (COG-UK) Consortium | Sergi Castellano, Rachel Williams, Mark Kristiansen, Paola Resende Silva, Sunando Roy, Tony Brooks, Helena Tutill, Paola Niola, Patricia Dyal, Charlotte Williams, Leysa Forrest, Yasmin Panchbhaya, Jacqueline Findlay, Samuel Weeks, Julianne Brown, Kathryn Harris, Paul Randell, James Price, Alison Holmes, Judith Breuer | EPI_ISL_839101 |

| | | | |
|---|---|---|---|
| Austrian Agency for Health and Food Safety (AGES) | Bergthaler laboratory, CeMM Research Center for Molecular Medicine of the Austrian Academy of Sciences | Lukas Endler, Anna Schedl, Fabian Amman, Petr Triska, Thomas Penz, Benedikt Agerer, Maelle Le Moing, Michael Schuster, Bekir Erguner, Jan Laine, Martin Senekowitsch, Christoph Bock, Andreas Bergthaler | EPI_ISL_2324194 |
| Department of Genetics, Medirex | Laboratory of Genomics and Bioinformatics, Comenius University Science Park | Tatiana Sedláčková, Miroslav Böhmer, Renáta Lukačková, Gabriel Minárik, Anna Gičová, Werner Krampl, Diana Rusňáková, Jaroslav Budiš, Tomáš Szemes | EPI_ISL_1280116, EPI_ISL_1280134, EPI_ISL_1280136, EPI_ISL_1280137, EPI_ISL_1280140, EPI_ISL_1280142, EPI_ISL_1280153 |
| Institute of Virology, Biomedical Research Center of the Slovak Academy of Sciences, Bratislava | Faculty of Natural Sciences, Comenius University, Bratislava | Broňa Brejová, Viktória Čabanová, Kristína Boršová, Viktória Hodorová, Sabina Fumačová Havlíková, Juraj Kopáček, Martina Ličková, Ľubomíra Lukáčiková, Martina Neboháčová, Monika Sláviková, Tomáš Vinař, Jozef Nosek, Boris Klempa | EPI_ISL_903986 |
| Austrian Agency for Health and Food Safety (AGES) | Bergthaler laboratory, CeMM Research Center for Molecular Medicine of the Austrian Academy of Sciences | Lukas Endler, Anna Schedl, Thomas Penz, Benedikt Agerer, Maelle Le Moing, Michael Schuster, Bekir Erguner, Jan Laine, Martin Senekowitsch, Christoph Bock, Andreas Bergthaler | EPI_ISL_1008144, EPI_ISL_1008146, EPI_ISL_1008163, EPI_ISL_1008172 |
| Lighthouse Lab in Milton Keynes | Wellcome Sanger Institute for the COVID-19 Genomics UK (COG-UK) Consortium | The Lighthouse Lab in Milton Keynes and Alex Alderton, Roberto Amato, Sonia Goncalves, Ewan Harrison, David K. Jackson, Ian Johnston, Dominic Kwiatkowski, Cordelia Langford, John Sillitoe on behalf of the Wellcome Sanger Institute COVID-19 Surveillance Team | EPI_ISL_720196, EPI_ISL_797130 |
| Eurofins LifeCodexx GmbH | Robert Koch Institute | unknown | EPI_ISL_1157070 |

| | | | |
|---|---|---|---|
| Institute of Virology, Biomedical Research Center of the Slovak Academy of Sciences, Bratislava | Faculty of Natural Sciences, Comenius University, Bratislava | Viktória Hodorová, Kristína Boršová, Broňa Brejová, Viktória Čabanová, Dominika Fričová, Sabina Fumačová Havlíková, Juraj Kopáček, Martina Ličková, Ľubomíra Lukáčiková, Martina Neboháčová, Monika Sláviková, Edita Staroňová, Elena Tichá, Tomáš Vinař, Jozef Nosek, Boris Klempa | EPI_ISL_577736, EPI_ISL_577738, EPI_ISL_583483 |
| Institute of Virology, Biomedical Research Center of the Slovak Academy of Sciences, Bratislava | Faculty of Natural Sciences, Comenius University, Bratislava | Kristína Boršová, Viktória Čabanová, Broňa Brejová, Viktória Hodorová, Sabina Fumačová Havlíková, Juraj Kopáček, Martina Ličková, Ľubomíra Lukáčiková, Martina Neboháčová, Monika Sláviková, Tomáš Vinař, Boris Klempa, Jozef Nosek | EPI_ISL_903990, EPI_ISL_788982 |
| Institute of Virology, Biomedical Research Center of the Slovak Academy of Sciences, Bratislava | Faculty of Natural Sciences, Comenius University, Bratislava | Broňa Brejová, Viktória Hodorová, Kristína Boršová, Viktória Čabanová, Dominika Fričová, Sabina Fumačová Havlíková, Juraj Kopáček, Martina Ličková, Ľubomíra Lukáčiková, Martina Neboháčová, Monika Sláviková, Edita Staroňová, Elena Tichá, Tomáš Vinař, Jozef Nosek, Boris Klempa | EPI_ISL_577740, EPI_ISL_577741, EPI_ISL_577742 |
| BTC, Khalifa University | BTC, Khalifa University | Al Safar et al | EPI_ISL_859852, EPI_ISL_859885 |
| Viollier AG | Department of Biosystems Science and Engineering, ETH Zürich | Chaoran Chen, Sarah Nadeau, Ivan Topolsky, Emmanouil Dermitzakis, Keith Harshman, Ioannis Xenarios, Henri Pegeot, Lorenzo Cerutti, Deborah Penet, Philipp Jablonski, Lara Fuhrmann, David Dreifuss, Katharina Jahn, Christiane Beckmann, Maurice Redondo, Olivier Kobel, Christoph Noppen, Sophie Seidel, Noemie Santamaria de Souza, Niko Beerenwinkel, Tanja Stadler | EPI_ISL_1004816, EPI_ISL_1004898, EPI_ISL_1004899, EPI_ISL_1119202 |

| | | | |
|---|---|---|---|
| Institute of Virology, Biomedical Research Center of the Slovak Academy of Sciences, Bratislava | Faculty of Natural Sciences, Comenius University, Bratislava | Kristína Boršová, Viktória Hodorová, Broňa Brejová, Viktória Čabanová, Dominika Fričová, Sabina Fumačová Havlíková, Juraj Kopáček, Martina Ličková, Ľubomíra Lukáčiková, Martina Neboháčová, Monika Sláviková, Edita Staroňová, Elena Tichá, Tomáš Vinař, Boris Klempa, Jozef Nosek | EPI_ISL_577735, EPI_ISL_577737, EPI_ISL_577739 |
| Department of Virus and Microbiological Special Diagnostics, Statens Serum Institut, Copenhagen, Denmark | Aalborg University | Danish Covid-19 Genome Consortium | EPI_ISL_1868624, EPI_ISL_1872494 |
| Institute of Virology, Biomedical Research Center of the Slovak Academy of Sciences, Bratislava | Faculty of Natural Sciences, Comenius University, Bratislava | Viktória Hodorová, Kristína Boršová, Broňa Brejová, Viktória Čabanová, Sabina Fumačová Havlíková, Juraj Kopáček, Martina Ličková, Ľubomíra Lukáčiková, Martina Neboháčová, Monika Sláviková, Tomáš Vinař, Jozef Nosek, Boris Klempa | EPI_ISL_718260 |
| Regional Authority of Public Health Banská Bystrica Slovakia | SVFI, Veterinary institute in Zvolen, Slovakia | Dirbáková Z., Maďarová L., Mancoš M., Strhársky J., Sujová S., Mokryšová S., Tinák M., Mojžiš M. | EPI_ISL_1942265 |
| Public Health Authority of the Slovak Republic | Bergthaler laboratory, CeMM Research Center for Molecular Medicine of the Austrian Academy of Sciences | Lukas Endler, Anna Schedl, Fabian Amman, Thomas Penz, Benedikt Agerer, Maelle Le Moing, Michael Schuster, Bekir Erguner, Jan Laine, Martin Senekowitsch, Christoph Bock, Andreas Bergthaler | EPI_ISL_1180701 |
| Labor Dr. Wisplinghoff - Köln | Robert Koch Institute, Bioinformatics MF1, Berlin, Germany | Dr. R. Grosser, Stephan Fuchs, Stefan Kroeger, Marianne Wedde, Oliver Drechsel, Aleksandar Radonic, Rene Kmiecinski, Ralf Duerrwald, Thorsten Wolff | EPI_ISL_751799 |
| Clinical Virology | Clinical Bacteriology | Tim Roloff, Madlen Stange, Helena MB Seth-Smith, Alfredo Mari, Karoline Leuzinger, Julia Bielicki, Manuel Battegay, Hans Hirsch, Adrian Egli | EPI_ISL_2270108 |

| | | | |
|---|---|---|---|
| Lighthouse Lab in Cambridge | Wellcome Sanger Institute for the COVID-19 Genomics UK (COG-UK) Consortium | Rob Howes, The Lighthouse Lab in Cambridge and Alex Alderton, Roberto Amato, Sonia Goncalves, Ewan Harrison, David K. Jackson, Ian Johnston, Dominic Kwiatkowski, Cordelia Langford, John Sillitoe on behalf of the Wellcome Sanger Institute COVID-19 Surveillance Team | EPI_ISL_719132, EPI_ISL_777815, EPI_ISL_777895 |
